# Determining the feasibility of linked claims and vaccination data for a Covid-vaccine pharmaco-epidemiological study in Germany – RiCO feasibility study protocol

**DOI:** 10.1101/2024.03.04.24303714

**Authors:** Nina Timmesfeld, Peter Ihle, Robin Denz, Katharina Meiszl, Katrin Scholz, Doris Oberle, Ursula Drechsel-Bäuerle, Brigitte Keller-Stanislawski, Hans H. Diebner, Ingo Meyer

## Abstract

In Germany, there has been no population-level pharmaco-epidemiological study on the safety of the Covid-19 vaccines. One factor preventing such a study so far relates to challenges combining the different relevant data bodies on vaccination with suitable outcome data, specifically statutory health insurance claims data. Individual identifiers used across these data bodies are of unknown quality and reliability for data linkage. As part of a larger pharmaco-vigilance study on the COVID-19 vaccines, called RiCO (German "Risikoevaluation der COVID-19-Impfstoffe”, Englisch "Risk assessment of COVID-19 vaccines”), a feasibility study is being conducted to determine the overall confidence level with which existing data can be analysed in relation to the safety of the COVID-19 vaccine. This RiCO feasibility study will establish a dataflow combining claims data and vaccination data for a sub-sample of the total German population, describe data quality for each data set from the various sources, estimate the proportion of the different linkage errors and will develop various approaches for linking the data in addition to the simple form of linkage using a common identifier in order to reduce possible linkage errors. These last three points are the core objective of the feasibility study. A secondary objective is to test the viability of the required dataflow involving multiple stakeholders from different parts of the healthcare system. Results will be published and used to plan the actual pharmaco-vigilance study on the COVID-19 vaccines for Germany, as well as future research on the role of COVID vaccines as risk or protective factors for long-term COVID-19 effects.

**Strength and limitations:**

- Potential for a population-level pharmaco-epidemiological study on the safety of the Covid-19 vaccines for Germany, based on vaccination data combined with statutory health insurance claims data.
- Introduction and estimation of quality metrics pertaining to the linkability of the various data bodies existing in Germany.
- Direct measurement of linkage error based on the available identifiers is not possible, proxy metrics and descriptive analytics need to be used.
- An attempt at linkage with the vaccination data could only be made using data from one smaller statutory health insurance, which may limit the extent to which the data can be analysed.

## Introduction

### Background

The World Health Organization (WHO) declared the SARS-CoV-2 pandemic on March 11, 2020 (1). To combat the COVID-19 pandemic, a large number of COVID-19 vaccines were developed and approved within a very short time (2). In parallel to the vaccination activities, in many countries pharmaco-epidemiological studies were conducted to investigate the safety of the different vaccines (3–6). In Germany, the RiCO study was initiated in 2020 as a population-level pharmaco-epidemiological study on the safety of the Covid-19 vaccines based on Covid 19 vaccination data and claims data from health insurance companies.

Until now, a lack of suitable data has hampered valid analyses. The reasons for this lie both in the structural characteristics of the healthcare system and in the way COVID-19 vaccination campaign was set up at the beginning of the pandemic. Structurally, there is no central, population-level database of relevant health data, such as diagnoses and treatments. The closest available data body comprises claims data held individually by the around 100 statutory and around 40 private health insurances, which is an issue of fragmentation. The German vaccination campaign was financed directly by the Ministry of Health up to April 2023, not using the usual reimbursement process via the insurances. Therefore, COVID-19 vaccination data for the initial period are not included in health insurance claims data, but form a separate data body, in turn split into various regional data sets. As a consequence of these conditions, the RiCO study could only achieve its objectives by linking multiple data sets from both data bodies on an individual level. This, in turn, was and has been and continues to be hampered by the lack of proven unique identifiers available in both data sets due to data protection concerns. There are structural weaknesses in the existing identifiers that can potentially lead to two types of errors in linking individuals across the datasets: false matches and complete or partial missing matches. An earlier simulation study done by our group (7) was able to show that complete missing matches do not lead to biased estimates in certain unadjusted designs, leaving false matches as well as partial missing matches in unadjusted designs and all types of linkage errors in adjusted designs as potential issues. However, the exact degree of uncertainty, is not known and cannot be derived a priori by logical means. It requires an actual linkage attempt and an a posteriori measurement of possible linkage errors.

### Objectives

Against this background, RiCO has initiated a feasibility study on data linkage to determine the level of confidence with which the existing data can be analysed with respect to RiCO’s primary objective around the safety of the COVID-19 vaccines. In case of successful conclusion, this will be followed by the full-fledged pharmaco-epidemiological study.

Specifically, the RiCO feasibility study will establish a dataflow bringing together claims data and vaccination data for a sub-sample of the whole population, describe data quality for each data set from the various sources, estimate the proportion of the different linkage errors and will develop various approaches to link the data in addition to the simple form of linkage using a common identifier in order to reduce possible linkage errors. These last three points are the core objective of the feasibility study. A secondary objective is to test the viability of the required dataflow involving multiple stakeholders from different parts of the healthcare system.

## Methods and analysis

### Data sources and data flow

The feasibility study will use two different data sources: (1) statutory health insurance claims data provided by the health insurance in combination with (2) COVID-19 vaccination data. The claims data are provided by one German statutory health insurance company for three years from 2019 to 2021. This corresponds to about 650,000 insured persons on average per year. Geographically, the health insurance company covers the whole of Germany with a regional focus on the Southwest. The insurance company was chosen based on its willingness to support the feasibility study and a sufficient number of insured persons and estimated vaccination cases (exposure) to be able to carry out the planned analyses. The COVID-19 vaccination data in principle cover the complete vaccination campaign for the German population. Both data sets are described in more detail below, as well as the data flow being established to pseudonymise both data sets on the basis of available identifiers, to bring them together in a secure analytic environment, to link them and to analyse them.

The claims data set [data set 1] is split into several tables or profiles. One profile contains demographic information on the insured persons, including gender, year of birth, place of residence (at time of data extraction), insurance periods and insurance status. Also included are diagnosis codes according to ICD-10-GM (8) from both primary care (GPs and specialists in an outpatient / practice setting, corresponding to German *ambulante Versorgung*) and secondary care (in the sense of hospital care, corresponding to German *stationäre Versorgung*). Hospital care data also include date of admission and discharge, type of admission and discharge (e.g. emergency admission), diagnoses at admission and discharge and codes for procedures performed. Finally included are data on medications prescribed in primary care, including on active ingredients, data of prescription and date of dispensing in the pharmacy.

The vaccination data [data set 2] are first separated based on the setting in which the vaccination was administered. The so-called DIM data set (corresponding to German *<u>D</u> igitales Impf <u>m</u> onitoring* or digital vaccination monitoring) contains data from vaccination centers, mobile vaccination teams (inter alia carrying out early vaccinations in nursing homes) and occupational physicians [data set 2a]. Data for this data set were collected centrally at German public health authorities Robert Koch Institute and Paul-Ehrlich-Institut. The so-called KV data set (corresponding to German *Kassenärztliche Vereinigung* or regional associations of statutory health insurance physicians) contains data from registered (primary care) physicians [data set 2b]. These data were first collected from primary care practices by the 17 regional associations and are currently being transferred to Robert Koch Institute and Paul-Ehrlich-Institut. Both variants of data set 2 have a similar data structure, including several identifiers (cf. below), day of vaccination, information on the vaccine administered and vaccination series (at least first, second or later vaccination for the individual). In the absence of any variable identifying the health insurance of the vaccinated person, the feasibility study uses a dataset of all vaccination cases to link to the claims data of the selected statutory health insurants.

The technical basis for data linkage is provided by the different identifiers contained in the three data sets 1, 2a and 2b. The starting point and weakest component in the entire approach is the set of identifiers in the DIM data set [2a]. These are the last name, first name and date of birth of the vaccinated person, as recorded in the vaccination centres etc. In the early stages of the vaccination campaign, methods of recording could include hand-written lists of names uttered by the vaccinated person to a member of the vaccination center team. Electronic means of recording were also used, as were automated registration processes using the electronic health insurance card (EHIC). The manner of recording can vary and is not documented with the data. These three identifiers were then used to generate a series of four consecutive pseudonyms (called ID1 to ID4) by hashing the concatenated identifiers (1=original values, 2=normed values, 3 and 4=using phonetical algorithms). Note that the mappings of ID1 to ID2 and from ID2 to ID3 and from ID2 to ID4 are unique mappings, but these are not always bijective, meaning that for example it may happen that two different ID1 are mapped to the same ID2. The Federal Press acted as a trust center for the pseudonymisation of all COVID-19 vaccination data. It is the only organisation holding the algorithms and keys (hash-function with salt-value) for this pseudonymisation process. Data set 2b (KV data set) contains the same identifier set ID1 to ID4. In addition, the physician associations collecting this data set generated a pseudonym based on the statutory health insurance number (referred to as identifier ID5) also hashed by the Federal Press software tool. According to available information, identifiers ID1 to ID4 in this data set were always based on names and date of birth taken from the EHICs of the insured persons. The health insurance number is a life-long unique identifier for all statutory health insurants. Data set 1 (claims data) contains the same identifier set as 2b, which were generated from Name, Surname and date of birth (ID1 to ID4) and the health insurance number (ID5) as they appear in the claims data.

Figure 1 depicts a simplified version of the data flow established for the RiCO feasibility study. It shows how the three constituent data sets 1, 2a and 2b are collected, processed and pseudonymised to build the amalgamated RiCO study data set.

**Figure 1.**
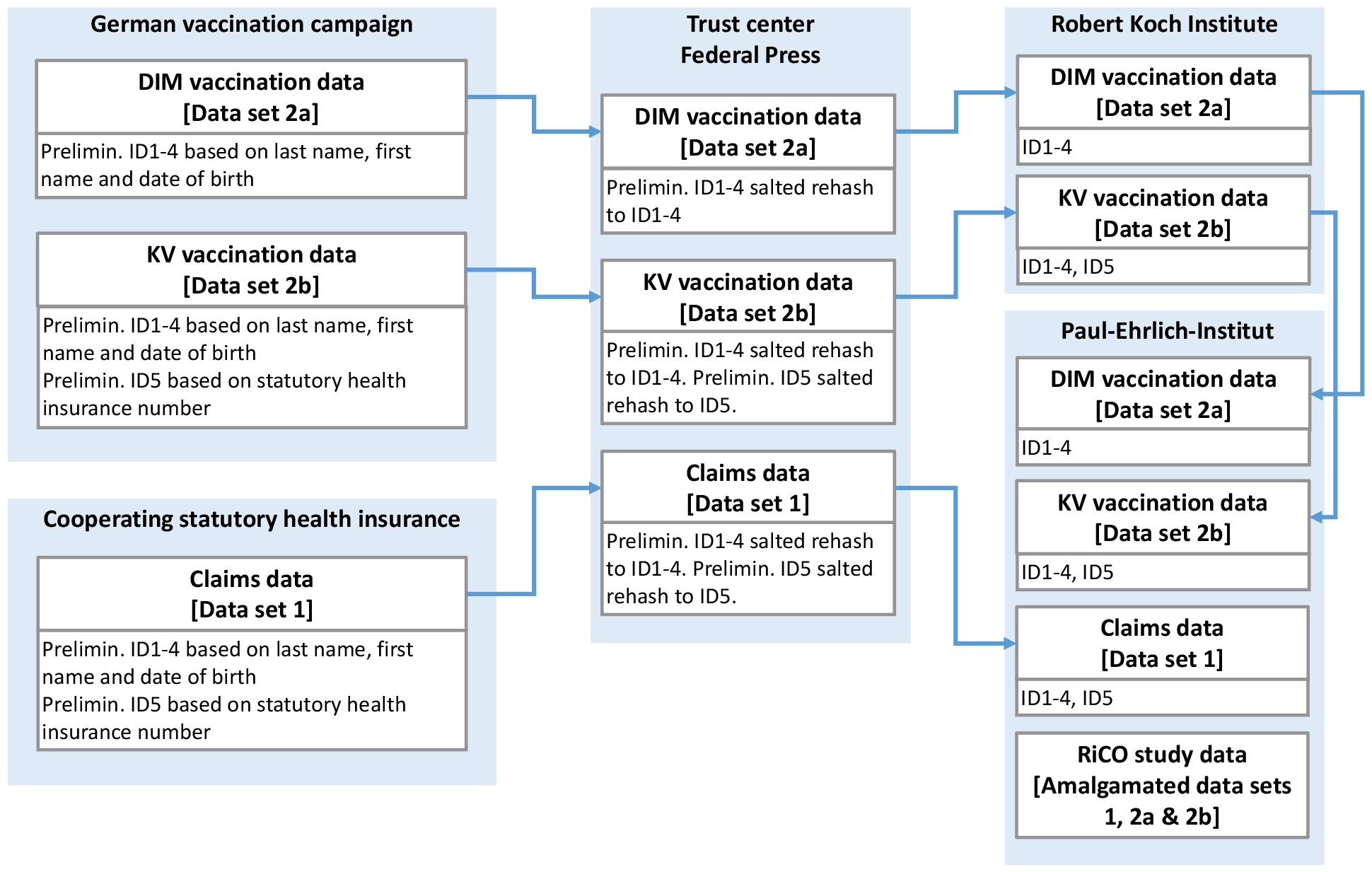
Simplified data flow RiCO feasibility study

### Data analysis

The RiCO study data set [amalgamated 1, 2a and 2b] is then analysed (1) in terms of data quality and (2) in terms of linkage quality.

#### Analysis of data quality

To describe data quality the following descriptive statistics will be calculated:

1. Proportion of duplicated rows in each of the data sets 2a and 2b.
2. For each vaccine, setting (DIM, KV) and the different age groups, proportion of coded vaccination dates set before the start of the vaccination campaign for this vaccine in the setting and age group.
3. Proportion of coded vaccination dates set after the closure of the vaccination centres in the respective federal states (only data set 2a).
4. Proportion of vaccination cases with identical ID1 but a least one difference in ID2 to ID4 in each of the data sets 2a and 2b.
5. Proportion of vaccination cases with missing ID5 in data set 2b.
6. Proportion of vaccination cases with identical ID5 but difference in ID1 and inconsistent age or sex in dataset 2b.
7. Proportion of incomplete vaccination series, e.g. only second vaccination for a person without first vaccination or missing second vaccination if third vaccination is available (data set with all vaccinations, 2a and 2b).
8. Proportion of vaccination series with implausible intervals between vaccinations (less than 2 weeks or greater than 13 weeks between the first and second or less than 5 months between all further vaccinations from the second vaccination onwards).

#### Analysis of linkage quality

In this setting, two kinds of linkage-errors are possible: *false matches and completely missing or partially missing matches*. False matches occur when the health-care data of one individual is mistakenly linked to the wrong persons Covid-19 vaccination information due to an identical pseudonym. This may happen if both individuals share all characteristics used to generate the pseudonym or if the identical characteristics is created due to spelling mistakes. Missing matches on the other hand occur when an individual received at least one Covid-19 vaccination, but the information about this vaccination could not be linked to the persons health-care data. This type of linkage error may occur, for example, when a name is misspelled in one of the data sources or has changed between the different times of data extraction. The category of missing matches may further be sub-divided into *completely missing matches* and *partially missing matches*. The former means that information about all vaccinations a person received could not be linked, while the latter entails cases in which only some of the Covid-19 vaccinations the person received failed to be linked with the health-care data.

All linkage errors can only occur during the linkage of claims data (data set 1) with the DIM data (data set 2a), because the KV data (data set 2b) contains the ID5 from the health insurance number, which is unique. However, all data sets can be used to estimate the amount of possible linkage errors. Obtaining reliable estimates of the amounts of linkage errors theoretically requires some “ground truth” data with record pairs that are known to be true matches (9). Due to administrative reasons, it was impossible to obtain such record pairs. However, there are some techniques that may still be used to assess the linkage quality, as discussed below.

##### False matches

These can occur due to the following reasons:

(FMR1) There exist two persons with the same name and date of birth.

(FMR2) Spelling mistakes resulting in two persons with similar names being merged into one, e.g. Lieschen Schmitt merges with Lieschen Schmidt.

(FMR3) As corrections for possible spelling mistakes are also made when generating the hashes for ID2-4, this can result in identical IDs despite different ID1 (name and date of birth).

The amount for the different reasons will be estimated from the different data sources as follows:

(FMR4) In the claims data (data set 1) as proportion with the same ID1-ID4 but different ID5, in the KV data (data set 2b) as proportion per vaccination series (only < 3) with the same ID1-ID4 and different ID5.

(FMR5) In the DIM data as proportion per vaccination series with the same ID1-ID4, which estimates the proportion due to reason (FMR1) or (FMR2)

(FMR6) Separately, in all data sets and for ID2-4 as proportion of same IDx but different ID1 per vaccination series (for data set 2b only < 3).

##### Missing matches

These can occur due to the following reasons:

(FMR7) Change of name between vaccinations or up to the time point of the data extraction from the statutory health insurance company.

(FMR8) Spelling mistakes resulting in the same person’s name diverging over time.

In both cases the ID1 from the DIM data (set 2a) did not match to the GKV data (set 1). Due to the spelling error correction when creating the hashes, it is possible that ID3 and 4 in particular are still correct in the second case.

The amount for the different reasons will be estimated from the different data sources as follows:

(FMR9) Separately from KV data and the linked KV and GKV data (linkage by ID5), rate of changes in ID1 with identical ID5 per month.

(FMR10) The size of the linkage error due to this reason cannot be estimated directly. It can also not be fully isolated from MMR1 (change of name) based on the available data. For both reasons, approximative quality metrics will instead be calculated as follows. First, the proportion of people (by ID1) with an incomplete vaccination series is estimated in the combined vaccination data set (2a and 2b, comprising the population of all vaccinated people across Germany) will be calculated. Second, for each month, age group and region, the following odds ratios will be estimated:

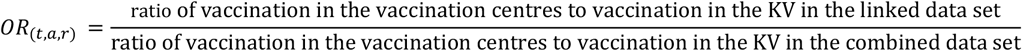

Both ratios are calculated from the number of vaccinations observed during the month (t) in this age group (a) and region (r) in the combined vaccination data set and the linked data set (data set 1 linked to combined vaccination data set using ID1, comprising all insurants of the participating statutory health insurance with a record in the vaccination data sets).

### Development of further linkage approaches

Since a reduction in the frequency of one linkage error always leads to an increase in the other, suggestions for the linkage procedure should be developed depending on the results, as well as on the subsequent analyses, e.g. cohort approach or self-controlled case series. Possible ideas include cleaning up the data, e.g. by removing duplicate IDs, taking into account other characteristics such as gender and place of residence, if necessary, as well as the vaccination serial number.

## Ethics and dissemination

In accordance with national legal requirements (§ 75 of Book X of the Social Security Code, SGBX), permission to use of the statutory health insurance claims data was requested and granted by the national regulatory authority for statutory health insurances. The vaccination data will be used in accordance with the requirements of § 3 of the national regulation on COVID-19 vaccinations (Verordnung zum Anspruch auf Schutzimpfung und auf Präexpositionsprophylaxe gegen COVID-19 (COVID-19-Vorsorgeverordnung). This regulatory framework permits the use of these pseudonymised data bodies without individual consent. Ethical approval was also not required for this secondary data study and was therefore not sought. All data processing is done in accordance with relevant legal and regulatory requirements in a secure computing environment hosted at Paul-Ehrlich-Institut.

After completion of the study, all data sets used will be archived for 10 years in accordance with good scientific practice.

The results of the feasibility study will be published in a suitable, peer-reviewed scientific journal. Results will also be presented to stakeholders from German statutory health insurances and government organisations.

## Discussion

Although the vaccination programme in Germany was launched more than three years ago, the data collected during the vaccination campaign have so far only been used to report vaccination rates. This is also due to the fact that not all data have been received by the two responsible federal authorities (PEI, RKI). For future scientific analyses, these vaccination data must be linked with other data sources such as claims data. This requires knowledge of the data quality, particularly the quality of the identifiers used with regard to possible data linkage.

Usually, the linkage quality is assessed relative to some “true values” (9). As this is not possible for the vaccination data collected in Germany, the aim of the planned feasibility study is to estimate the data quality based on the existing data sets, particularly with regard to the magnitude of possible linkage errors. This should enable future users of the data to assess which questions can be addressed by using the data.

Once the magnitude of the linkage errors has been estimated, proposals for a suitable process for data linkage for typical pharmaco-epidemiological study designs, such as self-control case series and cohort design, will be developed as part of the feasibility study. By extending the existing simulation study, the effects of linkage errors on bias and power will also be analysed. This will make it easier for future users of the data to assess the feasibility of their project idea in advance. This is particularly important, because linking the data to the claims data of the larger German health insurance companies would create one of the largest data sets for studying the efficacy and safety of the various Covid-19 vaccines. Such a data set would potentially also allow further research on long-term effects of COVID-19 and the possible role of vaccinations as risk or protective factors.

## Data Availability

The data used for the present study are not available for further use.

## Authors’ contributions

Prof. Dr. Nina Timmesfeld: Principal investigator, study objectives, study design, measured outcomes, data analysis plan

Dr. Peter Ihle: Study objectives, data sources and data flow, measured outcomes

Robin Denz: Measured outcomes, data analysis plan

Katharina Meiszl: Measured outcomes, data analysis plan

Dr. Katrin Scholz: Data sources and data flow

Dr. Doris Oberle: Study objectives, selection of ICD 10 codes, critical review of the manuscript

Ursula Drechsel-Bäuerle: Administrative support, critical review of the manuscript

Dr. Brigitte Keller-Stanislawski: Principal investigator, study design, study objectives

Dr. Hans H. Diebner: Measured outcomes, data analysis plan

Ingo Meyer: Principal investigator, study objectives, study design, legal requirements, ethics and dissemination

